# Temporal and Phylodynamic analyses of the 2026 Bundibugyo virus disease outbreak in the Democratic Republic of the Congo

**DOI:** 10.64898/2026.10.07.26365001

**Authors:** Tony Wawina-Bokalanga, Pierre Akilimali, Jonathan E. Pekar, Pascal Adroba-Tandele, Adrienne Amuri-Aziza, Eddy Kinganda-Lusamaki, Daan Jansen, Bernardo Gutierrez, Ciara Judge, Olga Ntumba-Tshitenge, Joshua Quick, Diallo Amadou Mouctar, Samuel Wilkinson, Jean-Claude Makangara-Cigolo, Marcel Lola-Loway, John O. Otshudiema, Nicksy Gumede, Félix Mulangu, Justus Nsio, Prince Akil-Bandali, Jepsy Yango, Dav M. Ebengo, Tania Bishola Tshitenge, Cris Kasita, Joelle Ngalula-Mudiayi, Mathias Mossoko, Pauline-Chloé Musuamba-Kayembe, Lievin Tibasima-Dhesa, Princesse Paku-Tshambu, Sabue Mulangu, Pius Z. Kabututu, Olivier Tshiani-Mbaya, Dieudonné M. Mvumbi, Marie-Roseline Belizaire, Anne W. Rimoin, Lorenzo Subissi, Martine Peeters, Ahidjo Ayouba, Mosoka Papa Fallah, Kevin K. Ariën, Sofonias K. Tessema, David Blazes, Nicholas J. Loman, Caroline Colijn, Olivier le Polain de Waroux, Joseph L-H. Tsui, Cathal Mills, Ellie Bourgikos, Samuel V. Scarpino, Áine O’Toole, Renny Doig, Guy Baele, Nohelia Navarrete, the Ebola outbreak response consortium, Christian Happi, Laurens Liesenborghs, Miles W. Carroll, Georges L. Mvumbi, Daniel Mukadi-Bamuleka, Piet Maes, Christian Ngandu, Jason Kindrachuk, Dieudonné Mwamba, Jean-Jacques Muyembe-Tamfum, Steve Ahuka-Mundeke, Koen Vercauteren, Moritz U.G. Kraemer, Andrew Rambaut, Placide Mbala-Kingebeni

**Affiliations:** Institut National de Recherche Biomédicale (INRB), Kinshasa, Democratic Republic of the Congo; Faculty of Medicine, University of Kinshasa (UNIKIN), Kinshasa, Democratic Republic of the Congo; Department of Clinical Sciences, Institute of Tropical Medicine, Antwerp, Belgium; Institut National de Santé Publique (INSP), Kinshasa, Democratic Republic of the Congo; Kinshasa School of Public Health, University of Kinshasa, Kinshasa, Democratic Republic of the Congo; Institute of Ecology and Evolution, University of Edinburgh, Edinburgh EH9 3FL, UK; Laboratoire Provincial de Santé Publique, Ituri, Democratic Republic of the Congo; TransVIHMI, Université de Montpellier, INSERM, IRD, Montpellier, France; Pandemic Sciences Institute, University of Oxford, Oxford, United Kingdom; World Health Organization Country Office, Kinshasa, Democratic Republic of the Congo; University of Birmingham, Birmingham, UK; Graduate School of Cellular and Biomedical Sciences, University of Bern, Bern, Switzerland; Institute of Social and Preventive Medicine, University of Bern, Bern, Switzerland; Division Provinciale de la Santé, Ituri, Democratic Republic of the Congo; World Health Organization Regional Office for Africa, Brazzaville, Republic of Congo; Direction Surveillance Epidémiologique, Ministry of Public Health, Hygiene and Social Welfare, Kinshasa, Democratic Republic of the Congo; Africa Centres for Disease Control and Prevention, Addis Ababa, Ethiopia; Department of Life Sciences, Faculty of Sciences and Technology, University of Kinshasa, Kinshasa, Democratic Republic of the Congo; Programme National de Lutte contre les Fièvres Hémorragiques Virales, Kinshasa, Democratic Republic of the Congo; Ridgeback Biotherapeutics, Miami, Florida, USA; Clinical Monitoring Research Program Directorate, Frederick National Laboratory for Cancer Research, Frederick, MD, USA; Department of Epidemiology, Jonathan and Karin Fielding School of Public Health, University of California, Los Angeles, CA, USA; World Health Organization, Geneva, Switzerland; Virology Unit, Department of Biomedical Sciences, Institute of Tropical Medicine, Antwerp, Belgium; Gates Foundation, Seattle, USA; Simon Fraser University, British Columbia, Canada; Institute for Experiential AI, Northeastern University, Boston, MA, USA; Department of Microbiology, Immunology and Transplantation, KU Leuven, Leuven, Belgium; Culmen International, Alexandria, VA, USA; African Center of Excellence for Genomics of Infectious Diseases, Redeemer’s University, Ede, Nigeria; Department of Biological Sciences, Faculty of Natural Sciences, Redeemer’s University, Ede, Nigeria; Rodolphe Merieux INRB-Goma Laboratory, Goma, Democratic Republic of the Congo; European Plotkin Institute for Vaccinology, Université Libre de Bruxelles (ULB), Brussels, Belgium; Department of Medical Microbiology & Infectious Diseases, Max Rady College of Medicine, University of Manitoba, Winnipeg, MB, Canada; South African National Bioinformatics Institute, University of the Western Cape, South Africa

**Author notes:** **Correspondence to:** Tony Wawina-Bokalanga, MD, PhD, Placide Mbala-Kingebeni, MD, PhD. Contributed equally as first authors (Tony Wawina-Bokalanga, Pierre Akilimali, Jonathan E. Pekar, Pascal Adroba-Tandele). Joint Senior Authors (Koen Vercauteren, Moritz U.G. Kraemer, Andrew Rambaut, Placide Mbala-Kingebeni). A complete list of authors and their affiliations is provided in the Supplementary Appendix.

## Abstract

The Democratic Republic of the Congo is experiencing a major outbreak of Ebola disease caused by *Orthoebolavirus bundibugyoense* (Bundibugyo virus), which was declared on May 15, 2026. To investigate the timing of emergence and early transmission, we generated near full-length Bundibugyo virus genomes from 686 PCR-confirmed individuals across 22 health zones. We estimated the time of emergence as late February 2026 (95% highest posterior density interval [HPDI]: January 21–March 25, 2026) and an evolutionary rate of 8.7×10^-4^ substitutions per site per year (95% HPDI: 7.6–9.9×10^-4^), with no evidence of mutations associated with rapidly growing lineages.

## INTRODUCTION

Ebola disease outbreaks continue to pose a substantial public health threat in the Democratic Republic of the Congo (DRC), where 17 outbreaks caused by two orthoebolavirus species, including *Orthoebolavirus bundibugyoense* (Bundibugyo virus, BDBV) and *Orthoebolavirus zairense,* have been detected since 1976^1,2^.

The 2026 Bundibugyo virus disease (BVD) outbreak was officially declared by the Ministry of Public Health, Hygiene and Social Welfare of the DRC on May 15, 2026^3^. The first cases of BVD were reported in Ituri province, the epicentre of the outbreak, and by October 5, 2026, the outbreak had spread to six additional provinces, some of which share international borders with the Central African Republic, South Sudan, and Uganda. As of October 5, 2026, the outbreak response coordination team of the DRC reported through National Public Health Institute (Institut National de Santé Publique (INSP)) 8,665 PCR-confirmed cases, including 4,178 deaths, across 64 of 167 health zones in seven provinces^4^.

This outbreak represents the second Ebola disease outbreak caused by BDBV in the DRC, and to date the second-largest Ebola disease outbreak, historically^1^. The scarcity of compatible molecular diagnostic assays in Ituri province, DRC, delayed the detection of BDBV and may have contributed to initial epidemiological growth. These diagnostic challenges, together with the lack of approved vaccines and therapeutics for BDBV underscores the urgent need to strengthen outbreak control measures and accelerate the development of effective medical countermeasures. Furthermore, the current outbreak is complicated by resource limitations, high population mobility and security challenges^5^.

Our initial analyses suggest that the 2026 BVD outbreak sparked by a new zoonotic spillover event^6^, but gaps remain in understanding of the timing of emergence, early growth patterns, and evolutionary rate^7^. To address these questions, we generated BDBV genomes from 686 PCR-confirmed individuals from 22 of the 55 health zones with documented cases between May and August 2026 and performed a temporal phylogenetic analysis to understand the evolutionary history and transmission dynamics of the early 2026 BVD outbreak.

## METHODS

### Samples from BDBV-infected individuals

Clinical specimens, including EDTA whole-blood samples from living patients and oropharyngeal swabs from deceased patients, were collected from suspected cases of BVD between May 1, 2026, and August 17, 2026, in Ituri and Nord-Kivu provinces, DRC. Specimens were collected as part of the public health emergency response and were therefore exempt from informed consent procedures. Permission to use de-identified samples, demographic and clinical data was granted by the ethics committee of the Kinshasa School of Public Health (ESP-UNIKIN, Kinshasa, DRC; approval number ESP/CE/81/2026).

### Viral genome sequencing

Samples were analyzed at the Provincial laboratory of Bunia, where the Institut National de Recherche Biomédicale (INRB) installed a lab dedicated to the BDBV outbreak response. Samples were tested using two molecular diagnostic platforms: the RealStar Filovirus Screen RT-PCR kit (Altona Diagnostics, Hamburg, Germany) and the RADIONE point-of-care (KH Medical, Pyeongtaek-si, South Korea). PCR-positive samples with cycle threshold (Ct) values <31 were selected and subsequently sequenced using a tiled-amplicon sequencing approach. A subset of samples collected between May 3 and 30, 2026 was sequenced using probe-based viral capture enrichment approach, as described by Aziza et al^7^. Viral genome sequences were derived from the RNA extracted from inactivated blood or oropharyngeal specimens. RNA were reverse transcribed into complementary DNA using the LunaScript RT SuperMix kit (New England Biolabs, Ipswich, MA, USA) and amplified by multiplex PCR using either pan-Ebola v2.0.0 or BDBV-specific v1.0.0 primer pools (https:artic.network/viruses/bdbv/bdbv-sequencing-guide.html). Sequencing libraries were prepared using the Native Barcoding Kit 96 V14 (SQK-NBD114.96; Oxford Nanopore Technologies (ONT)), following the manufacturer’s protocol. Negative controls were included throughout library preparation to monitor contamination. Final libraries were loaded on R10.4.1 flow cells (ONT, Oxford, UK) and run on a GridION sequencer. In addition, other PCR-positive samples from Ituri province were sequenced at INRB in Kinshasa using the pan-Ebola v2.0.0 primer pools. At INRB, sequencing libraries were prepared using either the ONT Native Barcoding Kit 96 V14 (SQK-NBD114.96) or the Illumina DNA Prep with enrichment (Illumina), following the manufacturer’s instructions. Libraries were loaded onto a GridION or an Illumina NextSeq 1000/2000 sequencer.

### Genomic data analysis

We generated 686 consensus genomes using either the ARTIC Network amplicon-nf Nextflow pipeline version 2.0 (https://github.com/artic-network/amplicon-nf/tree/0.4.6dev) for ONT long-reads or Viralrecon 2.6.0 (https://github.com/nf-core/viralrecon) for Illumina short-reads. Genomes were aligned to reference NC_014373.1 using MAFFT v7.525^8^ and the resulting alignment was subsequently trimmed to length 18,890 nucleotides to remove sequencing artefacts at the end of one genome (Sample ID 26FHV054, Pathoplexus ID: PP_006XHL9.3). We next used IQ-TREE v3.0.1^9^ to infer a maximum likelihood tree, with near-zero branches collapsed to zero, and perform ancestral state reconstruction. The best-fit model was identified using ModelFinder^10^ as TIM+F+R3, which we used in subsequent maximum likelihood phylogenetic inference.

We examined the genomes for signals of Adenosine Deaminase Acting on RNA (ADAR) editing type mutations, using raccoon (https://github.com/artic-network/raccoon) with default parameters, focusing on filtering T-to-C substitutions. After filtering, we used IQ-TREE 3 with the TIM+F+R3 model to root the tree using a best-fit rooting approach, identified and removed temporal outliers, and then reran IQ-TREE 3 to generate a final maximum likelihood tree.

To infer the timescale and basic epidemiological characteristics of the outbreak, we used BEAST X v10.6.0-beta4^11^, employing a SkyGrid non-parametric model^12^, a general time-reversible (GTR) substitution model^13^ with invariant sites, and a strict molecular clock model with a continuous-time Markov chain rate-reference prior on the clock model mean^14^. We examined the robustness of our results using datasets available earlier in the outbreak and both SkyGrid and exponential growth coalescent models. Detailed dry-laboratory protocols are provided in the Supplementary Appendix.

## RESULTS

Between May 1, 2026, and August 17, 2026, there were a total of 5,241 PCR-confirmed cases of BVD across 55 affected health zones, with a positivity rate of 41.3% among tested cases. Most cases were reported from Ituri province (n=4,439; 84.7%), specifically in the Bunia (n=1,191; 22.7%), Rwampara (n=862; 16.4%), Mongbwalu (n=575; 11.0%), and Nizi (n=553; 10.6%) health zones (Fig. 1A–B).

**Figure 1.**
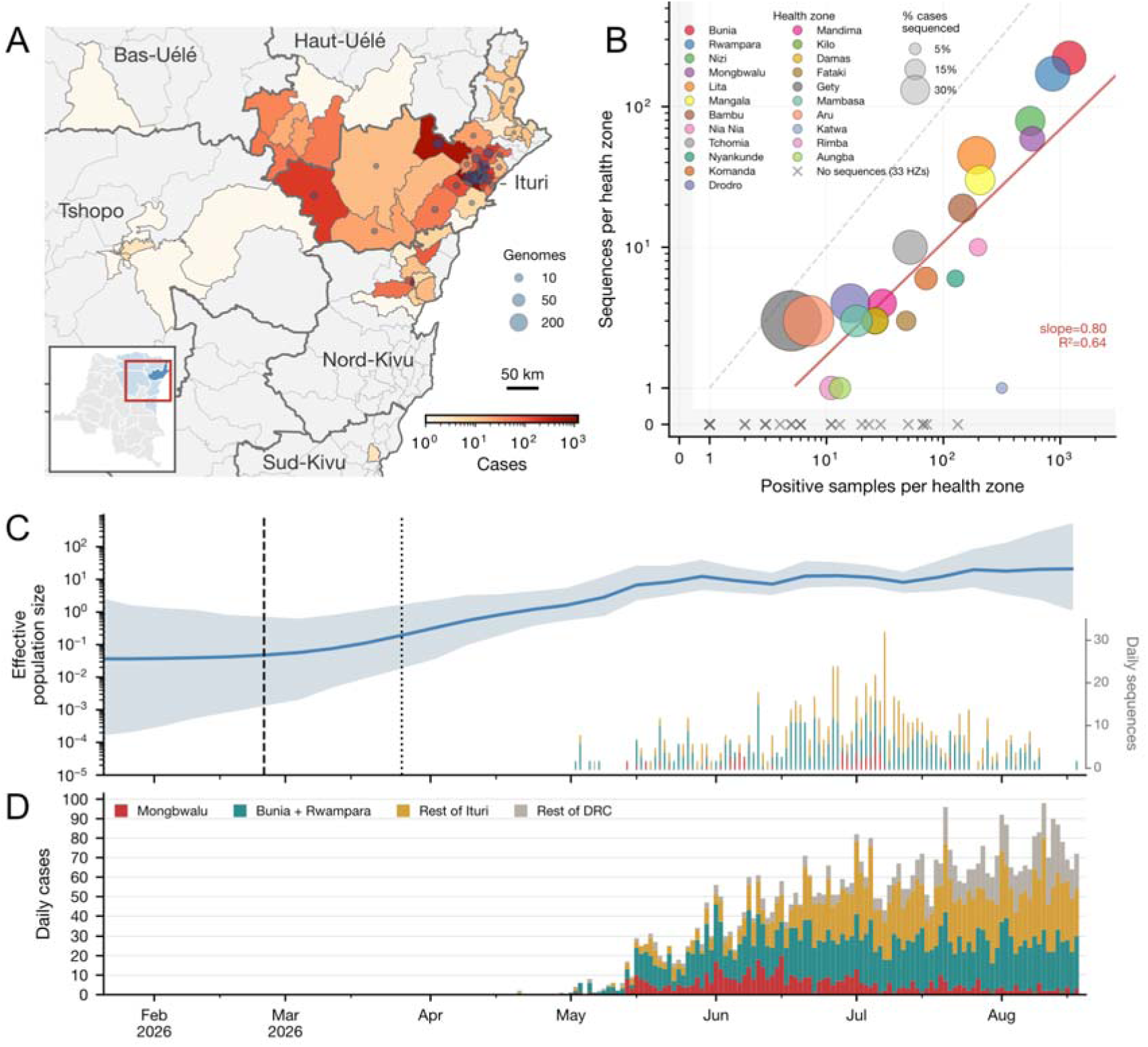
Epidemiological and evolutionary history of the early 2026 Bundibugyo virus disease outbreak in eastern DRC. **A,** Health zones with confirmed cases and genomes through August 17, 2026, with color of health zones and size of overlaid scatter indicating the number of cases and genomes, respectively. Inset indicates the affected provinces in blue, with Ituri in dark blue, with a red outline indicating the presented map. Labels indicate the six affected provinces. **B,** Genomic sampling intensity by health zone. Each circle represents a health zone, with size proportional to the percent of cases sequenced. The dashed grey unity line represents a sequence made available for every positive sample. The red line indicates the regression fit for health zones with at least one sequence, with slope and coefficient of variation (R^2^) shown. Shaded regions along each axis indicate zero values. Health zones with at least one positive sample but no sequences are indicated by an “x”. Health zones without any positive samples or sequences are excluded. **C,** Skygrid reconstruction of effective population size over time. The blue line shows the mean estimate, and the shaded region represents the 95% HPD interval. The dashed line indicates the median tMRCA; the dotted line indicates the upper bound of the 95% HPD interval; and the lower bound coincides with the lower limit of the y-axis (see Fig. S3 for a density plot of the tMRCA). A rug plot shows the dates and frequency of sample collection dates of genomes included in the analysis, colored as in panel D. **D,** Daily case counts (RT-qPCR positive) by symptom onset in Mongbwalu (red), Bunia and Rwampara (green), the rest of Ituri (gold), and the rest of the DRC (grey) (description of the data can be found in the Supplementary Appendix).

We generated near full-length BDBV genome sequences from 686 PCR-confirmed individuals, representing 13.1% of the reported confirmed cases of BVD in the DRC between May 1, 2026, and August 17, 2026 (Fig. 1B). Among the 22 health zones with available genome sequences, the number of genomes was associated with the number of PCR-positive samples per health zone (coefficient of variation R^2^ = 0.64, Fig. 1B).

The majority of sequences were from Ituri province and sequencing efforts only recently expanded to other provinces (Supplementary Appendix). Bunia (n=222), Rwampara (n=171), Nizi (n=79), Mongbwalu (n=59), Lita (n=45) and Mangala (n=30) health zones accounted for 606 (88.3%) of 686 genome sequences. The remaining 80 sequences were from 15 health zones in Ituri and 1 health zone in Nord-Kivu. The median age of BDBV-infected individuals sequenced was 28 years (IQR, 18–39; range, <1–84), and 353 (53.4%) of 661 samples with recorded sex were from females (Table 1; Table S1, and Fig. S1 in the Supplementary Appendix). Among samples for which the patient’s age was recorded (n = 607), children younger than 5 years accounted for 7.9% (n = 48) and individuals aged 18-44 years accounted for 58.1% (n = 353) (Table S2, and Fig. S1 in the Supplementary Appendix).

**Table 1.**
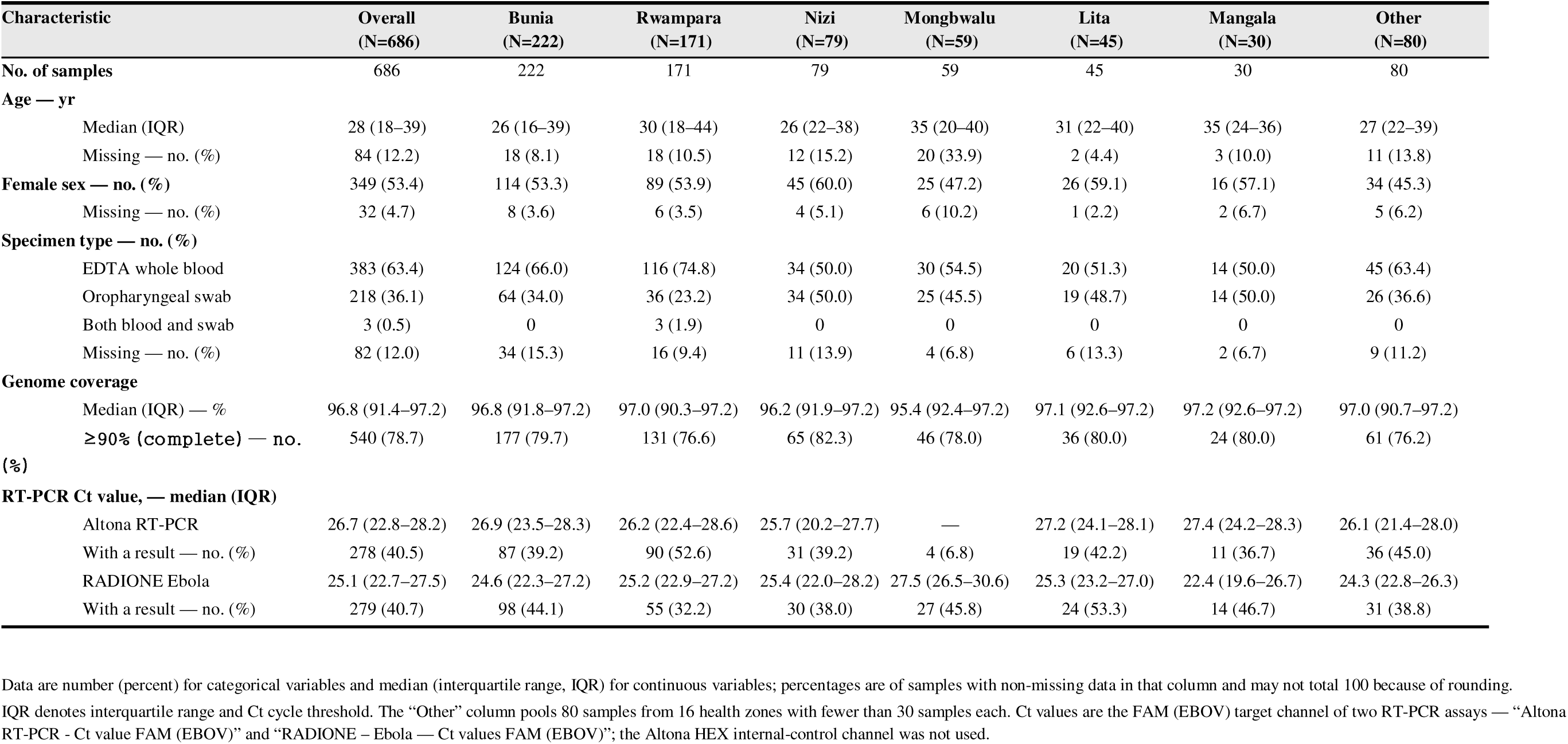
Characteristics of sequenced BDBV samples, overall and by health zone.

| Characteristic | Overall<br>(N=686) | Bunia<br>(N=222) | Rwampara<br>(N=171) | Nizi<br>(N=79) | Mongbwalu<br>(N=59) | Lita<br>(N=45) | Mangala<br>(N=30) | Other<br>(N=80) |
| --- | --- | --- | --- | --- | --- | --- | --- | --- |
| **No. of samples** | 686 | 222 | 171 | 79 | 59 | 45 | 30 | 80 |
| **Age — yr** |  |  |  |  |  |  |  |  |
| Median (IQR) | 28 (18–39) | 26 (16–39) | 30 (18–44) | 26 (22–38) | 35 (20–40) | 31 (22–40) | 35 (24–36) | 27 (22–39) |
| Missing — no. (%) | 84 (12.2) | 18 (8.1) | 18 (10.5) | 12 (15.2) | 20 (33.9) | 2 (4.4) | 3 (10.0) | 11 (13.8) |
| **Female sex — no. (%)** | 349 (53.4) | 114 (53.3) | 89 (53.9) | 45 (60.0) | 25 (47.2) | 26 (59.1) | 16 (57.1) | 34 (45.3) |
| Missing — no. (%) | 32 (4.7) | 8 (3.6) | 6 (3.5) | 4 (5.1) | 6 (10.2) | 1 (2.2) | 2 (6.7) | 5 (6.2) |
| **Specimen type — no. (%)** |  |  |  |  |  |  |  |  |
| EDTA whole blood | 383 (63.4) | 124 (66.0) | 116 (74.8) | 34 (50.0) | 30 (54.5) | 20 (51.3) | 14 (50.0) | 45 (63.4) |
| Oropharyngeal swab | 218 (36.1) | 64 (34.0) | 36 (23.2) | 34 (50.0) | 25 (45.5) | 19 (48.7) | 14 (50.0) | 26 (36.6) |
| Both blood and swab | 3 (0.5) | 0 | 3 (1.9) | 0 | 0 | 0 | 0 | 0 |
| Missing — no. (%) | 82 (12.0) | 34 (15.3) | 16 (9.4) | 11 (13.9) | 4 (6.8) | 6 (13.3) | 2 (6.7) | 9 (11.2) |
| **Genome coverage** |  |  |  |  |  |  |  |  |
| Median (IQR) — % | 96.8 (91.4–97.2) | 96.8 (91.8–97.2) | 97.0 (90.3–97.2) | 96.2 (91.9–97.2) | 95.4 (92.4–97.2) | 97.1 (92.6–97.2) | 97.2 (92.6–97.2) | 97.0 (90.7–97.2) |
| ≥ 90% (complete) — no. (%) | 540 (78.7) | 177 (79.7) | 131 (76.6) | 65 (82.3) | 46 (78.0) | 36 (80.0) | 24 (80.0) | 61 (76.2) |
| **RT-PCR Ct value, — median (IQR)** |  |  |  |  |  |  |  |  |
| Altona RT-PCR | 26.7 (22.8–28.2) | 26.9 (23.5–28.3) | 26.2 (22.4–28.6) | 25.7 (20.2–27.7) | — | 27.2 (24.1–28.1) | 27.4 (24.2–28.3) | 26.1 (21.4–28.0) |
| With a result — no. (%) | 278 (40.5) | 87 (39.2) | 90 (52.6) | 31 (39.2) | 4 (6.8) | 19 (42.2) | 11 (36.7) | 36 (45.0) |
| RADIONE Ebola | 25.1 (22.7–27.5) | 24.6 (22.3–27.2) | 25.2 (22.9–27.2) | 25.4 (22.0–28.2) | 27.5 (26.5–30.6) | 25.3 (23.2–27.0) | 22.4 (19.6–26.7) | 24.3 (22.8–26.3) |
| With a result — no. (%) | 279 (40.7) | 98 (44.1) | 55 (32.2) | 30 (38.0) | 27 (45.8) | 24 (53.3) | 14 (46.7) | 31 (38.8) |

Data are number (percent) for categorical variables and median (interquartile range, IQR) for continuous variables; percentages are of samples with non-missing data in that column and may not total 100 because of rounding. IQR denotes interquartile range and Ct cycle threshold. The “Other” column pools 80 samples from 16 health zones with fewer than 30 samples each. Ct values are the FAM (EBOV) target channel of two RT-PCR assays — “Altona RT-PCR - Ct value FAM (EBOV)” and “RADIONE – Ebola — Ct values FAM (EBOV)”; the Altona HEX internal-control channel was not used.

Using a SkyGrid coalescent model, we inferred an evolutionary rate of 8.7×10^-4^ substitutions per site per year (95% highest posterior density interval [HPDI]: 7.6–9.9×10^-4^), with a time of the most recent common ancestor (tMRCA) of late February 2026 (95% HPDI: January 21 – March 25, 2026; Fig. 1C). We find that the epidemic continued to grow between its emergence and the end of the study period (August 17, 2026) in line with epidemiological data (Fig. 1C, and Fig. 1D). We estimate a time-varying rate of growth with a relatively slower start in January and February 2026, an acceleration between March and June 2026, and a subsequent decline in growth and stabilization of the effective population size. The subsequent decline in growth rate pattern was reproduced using multiple data subsets with different cut-off dates, indicating that our modeling framework is robust to temporal sampling bias (Fig. 2).

**Figure 2.**
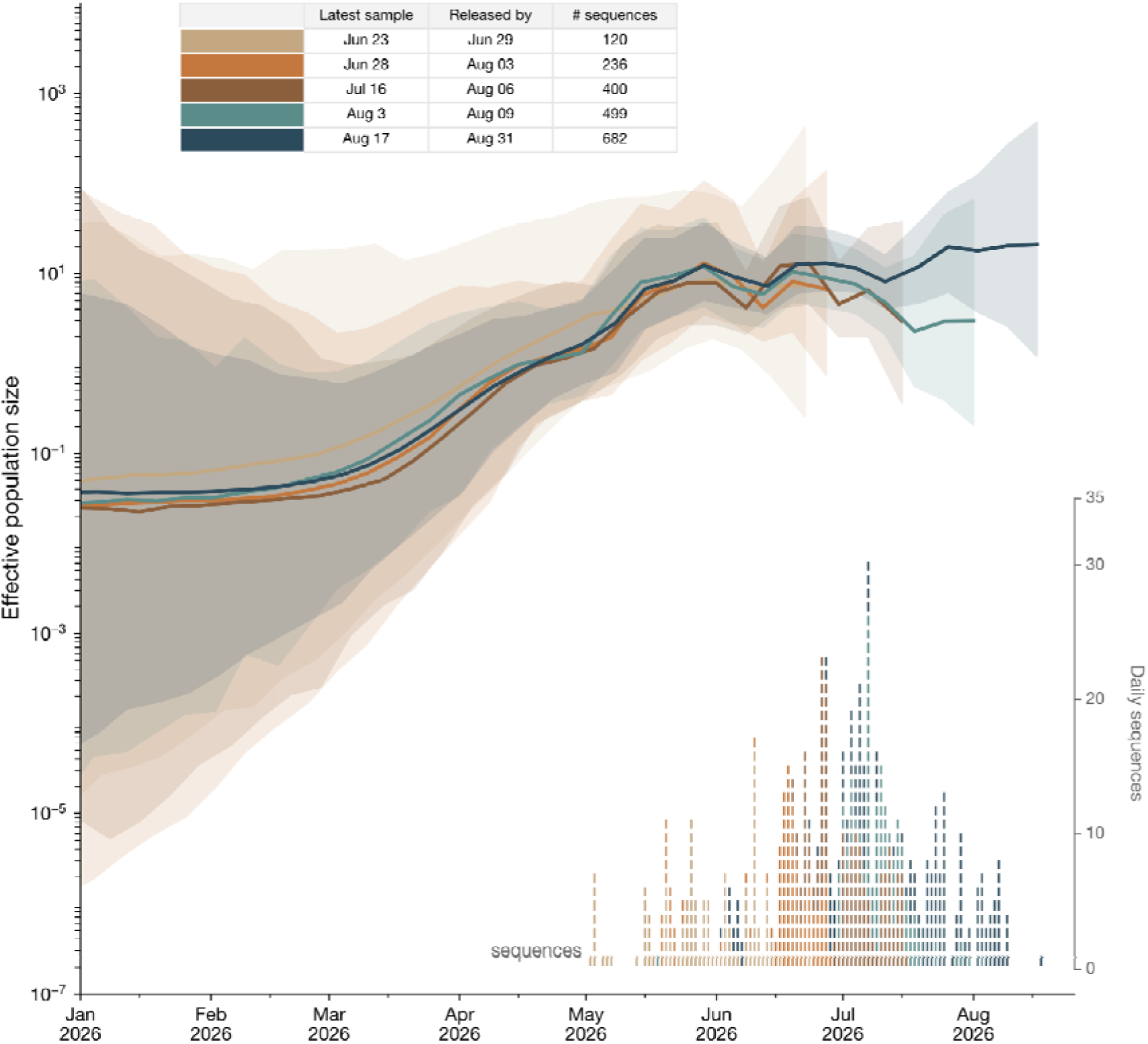
SkyGrid reconstruction of effective population size, when examining genomes available by a given release date. The solid line shows the median estimate, and the shaded region represents the 95% HPDI. Each dataset is colored by release date, with the most recent sample date and release date indicated in the legend. A rug plot shows the dates and frequency of collection dates of genomes included in the analysis, colored by which dataset first includes them. Each dataset shows an increase in both the temporal range and sampling depth. All analyses show an increase in the growth rate between March and June 2026, followed by a slight decline and stabilization. For the earliest cutoff with the fewest genomes, the period of stabilization is shorter, reflecting the earlier sampling cutoff. The analyses with cutoffs of 16 July and 3 August show a small decline toward the end of the sampling period, but these declines are brief and likely artifacts of sparse sampling near the window edge rather than genuine changes in the effective population size. The broader stabilization pattern is visible across all datasets, suggesting that once sufficient genomic diversity is available, phylodynamic analyses can reliably reconstruct the outbreak history and complement case-based surveillance.

We estimate a growth rate of 0.052 days^-1^ (95% HPDI: 0.025–0.084) and a corresponding doubling time of 13.2 days (95% HPDI: 7.1–23.7) before mid-June, when the SkyGrid reconstruction indicates approximately exponential growth (Fig. 1C, and Fig. S5 in the Supplementary Appendix). Applying an exponential model to the full dataset, however, yields a worse fit with an attenuated growth rate, as the estimate is averaged across the subsequent decline in growth (Fig. S5).

Genomes from Mongbwalu appear to represent substantial diversity in the maximum likelihood phylogeny, as well as emerging from relatively deep parts of both the maximum likelihood and time-calibrated phylogenies (Fig. S3 and Fig. S4 in the Supplementary Appendix). The root age and Skygrid reconstruction are compatible with the slow start of the epidemic in Mongbwalu, followed by exportations and an increased growth rate as larger urban areas (*e.g.*, Bunia) were seeded. These results would be concordant with epidemiological reports suggesting that Mongbwalu could have been a starting point for the outbreak^3^ and an observed recent decline in the daily number of new cases in Mongbwalu (Fig. 1D).

We identified 767 nucleotide variants within the outbreak dataset, most of which were unique to individual genomes: 62.1% occurred in a single genome and 82.7% in ≤3 genomes (Table S4, and Fig. S6). Two nucleotide variants nevertheless exceeded 10% prevalence, including one intergenic change (A10319G) and one missense substitution in the nucleoprotein (Y472D). One genome also carried the A82V mutation in the glycoprotein, previously linked to increased viral entry efficiency^15^. Importantly, we observed a low genetic diversity within the 2026 BVD outbreak, with no evidence that a lineage had a fitness advantage.

## DISCUSSION

On May 17 and 18, 2026, the World Health Organization and Africa Centres for Disease Control and Prevention (Africa CDC) declared the 2026 BVD outbreaks in the DRC and Uganda as a Public Health Emergency of International Concern^16^ and a Public Health Emergency of Continental Security^17^, respectively. Within four months of the 2026 BVD outbreak declaration, the reported cases have already far exceeded the DRC 2018-2020 Ebola virus disease outbreak, which resulted in 3,470 cases and 2,287 deaths^1^.

Our molecular dating analysis places the origin of the currently sampled BDBV diversity between mid-January and March 2026. This is consistent with reports of clinically compatible cases of Ebola disease in Mongbwalu in January 2026, suggesting that unsampled transmission may have preceded the diversity captured in this outbreak and the origin of the outbreak could be earlier^18^. However, our characterization of the general trajectory of the outbreak is robust to sampling cutoffs, indicating that early outbreak dynamics can be confidently estimated from genomic data once sufficient diversity is available, and that genomic analyses can serve as an independent and complementary data stream to epidemiological surveillance.

With genomes from Mongbwalu distributed across deep branches in the phylogeny, but the majority of genomes sampled through mid-August 2026 from Bunia and Rwampara, it is plausible that Mongbwalu was an early source of virus lineages that seeded these larger areas. An expansion to Bunia and Rwampara, and subsequent exponential growth would be consistent with epidemiological reports and expectations of pathogens spreading more rapidly in dense urban areas. As genomes from other health zones do not consistently form distinct clusters, there may have been frequent lineage movement during the rapid growth phase of the outbreak, consistent with documented human mobility patterns in the region^19^.

Nearly all samples analysed here are from Ituri, and additional genome sequencing is underway in other affected provinces (e.g., Nord Kivu and Haut-Uélé). Linking viral genomic and contact tracing data, alongside continued analysis of the temporal and geographic clustering of cases, could help identify spatial movements and their drivers, thereby guiding the ongoing outbreak response. Although we did not identify mutations associated with rapidly growing BDBV lineages, continued near-real time genomic surveillance is warranted, particularly in light of the ongoing deployment of medical countermeasures, including antivirals and vaccines.

## Data and materials availability

The genome sequences can be accessed as Pathoplexus SeqSet BDBV_DRC_20260831, are available at https://doi.org/10.62599/PP_SS_4291.1, and should be appropriately cited in any publications that use these data. Use of the data is governed by the Pathoplexus Data Use Terms: https://pathoplexus.org/about/terms-of-use/data-use-terms. All data and code to reproduce the results will be available on publication.

## Supporting information

Figure S3. Maximum likelihood phylogeny of the 2026 BVD outbreak in eastern DRC.

Figure S4. Time-resolved phylogeny of the 2026 BDBV outbreak in the DRC.

Figure S5. Comparison of exponential growth and SkyGrid coalescent models using the earliest release subsample (left; n=120, sampled through 23 June 2

## Supplementary Appendix

## Ebola outbreak response consortium

**Institut National de Recherche Biomédicale (INRB), Kinshasa, DRC**: Rilia Ola-Mpumbe, Gradi Luakanda-Ndelemo, Servet Kimbonza, Fiston Cikaya-Kankolongo, Tina Bakutumba-Loweya, Emmanuel Lokilo-Lofiko, Sifa Kavira-Bapitani, David Isengelo-Sikatenda, Judith Tete-Sitra, Andre Citenga, Julie Tuenakoko-Kulumbula, Nelson Kashali, Baby Muyembe, Kelly-Michel Kenye.

**Laboratoire Provincial de Santé Publique, Ituri, DRC**: Phine Malengo-Olokakoy, François Berocan-Underos, Berkias Bakambu-Nebape, Neema Kavugho-Sindani.

**Rodolphe Merieux INRB-Goma Laboratory, Goma, DRC:** Noella Mulopo-Mukanya, Brigitte Modadra-Madakpa, Michel Ngimba.

**Ministry of Public Health, Hygiene and Social Welfare, Kinshasa, DRC:** Jean-Marie Ndjemba-Yermbangh, Leonie Manya-Kitoto, José Cilumbayi.

**Culmen International, LCC:** Francis M. Mbuyi, Vandersal-Alain Salaboni, Gerry Vediena Makaya, Cristel Flores.

## Author contributions

Conceptualisation and study design: T.W.B., J.J.M.T., M.U.G.K., A.R., and P.M.B. Fieldwork supervision: T.W.B., P.A., P.A.T., S.A.M., and P.M.K. Sequencing: T.W.B., A.A.A., P.A.B., and P.C.M.K. Data curation: T.W.B., E.K-L., J.P., A.R., B.G., C.J., M.UG.K. Data visualisation: J.E.P. and M.U.G.K. Bioinformatics analysis: T.W.B., A.A.A., E.K.L., D.J., J.Q., S.W., N.J.L, and P.P.T. SNP analysis: D.J. Phylogenomic analyses: J.E.P. and A.R. Writing, first draft: T.W.B., J.E.P., and M.U.G.K. All authors reviewed and approved the final version of the manuscript.

## Acknowledgments

The authors are grateful to all staff involved in the Ebola disease outbreak response and the Ministry of Public Health, Hygiene and Social Welfare of the DRC. We acknowledge support from the World Health Organization, the Africa Centers for Disease Control, the UN Humanitarian Air Service, and the Bill & Melinda Gates Foundation. Sequencing activities were supported in part by the University of Manitoba (grant no. MRR-184813, to Jason Kindrachuk), the Institute of Tropical Medicine (ITM, Antwerp) through Belgian Directorate-general for Development Cooperation and Humanitarian Aid (DGD FA5 project), the Culmen International LCC, the Agence Française de Développement through the AFROSCREEN project (grant no. CZZ3209) coordinated by ANRS Maladies Infectieuses Emergentes in partnership with Institut Pasteur and Institut de Recherche pour le Développement (IRD), and the French Ministry of Europe and Foreign Affairs through the FEF programme. We also acknowledge support from the European Union (DECIPHER project under the Horizon Europe research and innovation program, and the grant agreement no. 101137242). Views and opinions expressed are those of the authors and do not necessarily reflect those of the EU or the granting authority EU Horizon Europe research and innovation program. The EU or the granting authority cannot be held responsible for them. J.Q., S.W., N.L., A.o.T., P.M. and A.R acknowledge the support of the Wellcome Trust through the ARTIC Network (awards 206298/Z/17/Z and 313694/Z/24/Z). M.U.G.K. acknowledges funding from The Rockefeller Foundation (PC-2022-POP-005), Health AI Programme and Collaborative in Disease Surveillance from Google.org, the Oxford Martin School Programmes in Pandemic Genomics & Digital Pandemic Preparedness, European Union’s Horizon Europe programme projects MOOD (#874850) and E4Warning (#101086640), Wellcome Trust grants 303666/Z/23/Z, 226052/Z/22/Z & 228186/Z/23/Z (The Global.health Project), the United Kingdom Research and Innovation (#APP8583), the Medical Research Foundation (MRF-RG-ICCH-2022-100069), UK International Development (301542-403), the Bill & Melinda Gates Foundation grants (INV-063472, INV-090281, INV-103122) and Novo Nordisk Foundation (NNF24OC0094346). A.A.-A. is supported by a DGD sandwich PhD scholarship, and D.J., a postdoctoral researcher, is supported by a FWO (Research Foundation Flanders) scholarship (grant number: 12AKD26N)

## Methodology

### Genomic data analysis

We examined the genomes for signals of Adenosine Deaminase Acting on RNA (ADAR) editing type mutations (*i.e.*, clusters of T-to-C substitutions), using raccoon (https://github.com/artic-network/raccoon) with default parameters: a 300-nucleotide window and minimum of 3 A-to-G and/or T-to-C substitutions to flag ADAR-editing. We focused on filtering T-to-C substitutions and left the A-to-G substitutions because they were relatively uncommon. Only one branch flagged by raccoon exhibited 3 A-to-G substitutions; all other branches flagged by raccoon only had 1 A-to-G substitution, with the rest being T-to-C substitutions. Because the glycoprotein exhibited excess T-to-C substitutions (n=24; Table S3), we masked these sites across all genomes in the alignment. After accounting for the ADAR-edited sites in the glycoprotein (GP) and discounting the one branch that only exhibited A-to-G substitutions, there were 8 tip branches and 3 internal branches that exhibited ADAR-editing signatures. Among these, we masked all but one ADAR-edited site, but in this case, only among the genomes that were affected by the ADAR edits (Table S3).

We reran IQ-TREE 3^1^ using the best-fit model, rooted the tree using a best-fit rooting approach that minimizes the residual sum of squares from a root-to-tip regression, and then identified temporal outliers as genomes outside of ±2 standard deviations of the residuals from the root-to-tip regression. The tree was rerooted whilst excluding the outliers; the outliers were added back in; the residuals were recalculated using the new root; and the process was repeated until the root stabilized. This iterative process eventually identified 35 outliers, from which we only removed four that had a z-score greater than 3, resulting in a final alignment of 682 genomes with 681 from Ituri and 1 from Nord Kivu. We then used IQ-TREE 3 once more to generate the final maximum likelihood tree.

To infer the timescale and basic epidemiological characteristics of the outbreak, we used BEAST X v10.6.0-beta4^2^, employing a SkyGrid non-parametric model^3^ with 39 transition points at one-week intervals and a cutoff of 40 weeks, a general time-reversible (GTR) substitution model^4^ and a strict molecular clock model with a continuous-time Markov chain (CTMC) rate-reference prior on the clock model mean^5^. A subset of genomes did not have sampling dates available; for these we used the date the sample was received for sequencing, as samples were typically received within 1–2 days of collection.

We ran an MCMC analysis of 100 million generations, subsampling every 10 thousand iterations to continuous parameter log and tree files.

To examine whether datasets available earlier in the outbreak would have yielded similar inferences, we created cumulative subsets of the 682 genomes based on their release date, using cutoffs of 1 July (n=120), 3 August (n=236), 6 August (n=400), and 10 August 2026 (n=499). We ran BEAST X with the same parameterizations as above, except adjusting the number of intervals for the SkyGrid model to match the time span of each subset. We additionally ran BEAST X with an exponential growth model on the earliest dataset and the complete dataset, with the remaining parameters the same as described above.

To reconstruct the ancestral sequence of the 2026 BDBV outbreak, we employed BEAST X with a GTR substitution model, a strict molecular clock model with a CTMC prior on the clock model mean, and an empirical tree distribution from our primary analysis. For this analysis, we used the same 682 taxa from the primary analysis, but in this case we retained all ADAR-edited sites to preserve the complete sequence for ancestral reconstruction. We ran an MCMC analysis of 100 thousand generations, subsampling every 10 iterations to continuous parameter log files. After discarding a 10% burn-in, site-wise nucleotide posteriors were summarized from the remaining MCMC samples. At each site, the ancestral base was called if its posterior exceeded 0.8. Otherwise the site was coded as ambiguous (N). The reconstructed ancestral sequence was then incorporated into the alignment and mutations in each of the 682 outbreak genomes were identified relative to this ancestral sequence and classified as intergenic, sense, missense or nonsense (Table S4).

Convergence and mixing for each analysis were assessed in Tracer v1.7.2^6^, and 10% of the chain was discarded as burn-in. All relevant effective sample size (ESS) values were >150.

All phylogenetic trees were visualized using Baltic (https://github.com/evogytis/baltic).

#### Missing genomic surveillance

There were 33 health zones with PCR-confirmed cases but without any sequenced genomes, representing Nord Kivu (n=11 health zones with confirmed cases but no sequenced genomes), Tshopo (n=7), Ituri (n=7), Haut-Uélé (n=6), Bas-Uélé (n=1), and Sud Kivu (n=1), indicating substantial lack of genomic surveillance outside of Ituri province and in lower-incidence health zones within it.

#### Ct value variation

Ct also varied by specimen type: On Altona the median Ct was 0.6 cycles lower in swabs than whole blood (relative difference −2.0%; 95% CI −8.1 to 3.2), and on RADIONE the difference was of similar magnitude (−0.6 cycles; −2.3%; 95% CI −9.1 to 4.2); on both platforms the CI included zero (Table S2).

#### Sample details

Data were missing for age in 84 samples (12.2%), sex in 32 (4.7%) and specimen type in 82 (12.0%). Genome coverage was high (median, 96.8%; IQR, 91.4–97.2), with 540 samples (78.7%) reaching ≥90%. Ct values from the FAM (EBOV) channel of the Altona RT-PCR and RADIONE Ebola assays were available for 278 (40.5%) and 279 (40.7%) samples, respectively (24 had a result on both); the median Ct was 26.7 (IQR, 22.8–28.2) for Altona and 25.1 (IQR, 22.7–27.5) for RADIONE. Higher Ct (lower viral load) was associated with modestly lower genome coverage, more so for Altona (Spearman ρ=-0.222, n=278) than for RADIONE (ρ=-0.132, n=279) but coverage remained high across the observed Ct range (Fig. S2).

#### Epidemiological data

We extracted the total number of cases by health zone reported between 1st of May and August 17th from the Situation Reports published by the Institut National de Santé Publique (INSP)^7^. Time series of confirmed and negative cases at the health zone level were extracted from the epidemiological line list, a central register of individual case reports across health care facilities in the affected provinces. Only reports of confirmed cases (i.e., records with a PCR-positive result) with symptom onset dated up to August 17, 2026, were included and aggregated to the health zone level. One positive case from the Viadana health zone was excluded from downstream analyses because there were no confirmed cases in Viadana published by the INSP by August 17, 2026. For records with a positive test result and no reported date of symptom onset but a known sample collection date, we impute the symptom onset dates based on fitted distributions between the onset to sample collection date (using the epidist package in R (version 0.4) as described by Ebengo et al^8^.

Health zone-level case counts displayed in the map (Fig. 1A) are derived from the INSP weekly situation reports, which provide cumulative confirmed case counts by health zone. All other analyses use the epidemiological line list described above. Minor discrepancies between the two sources reflect differences in case validation timelines. Data are regularly updated and presented on a dashboard^9^.

**Figure S1.**
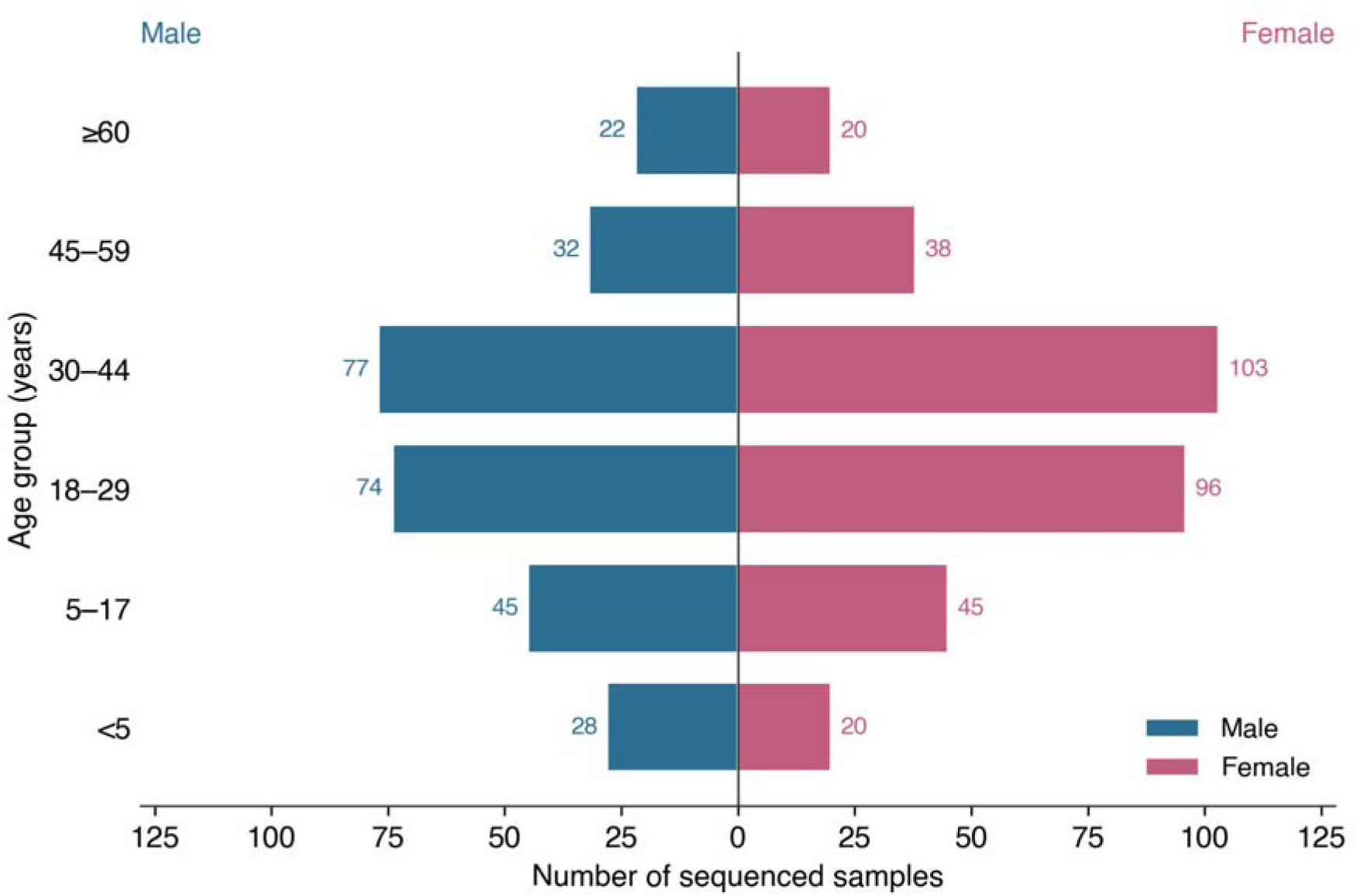
Age–sex distribution of individuals with sequenced samples (population pyramid). Blue, male; red, female.

**Figure S2.**
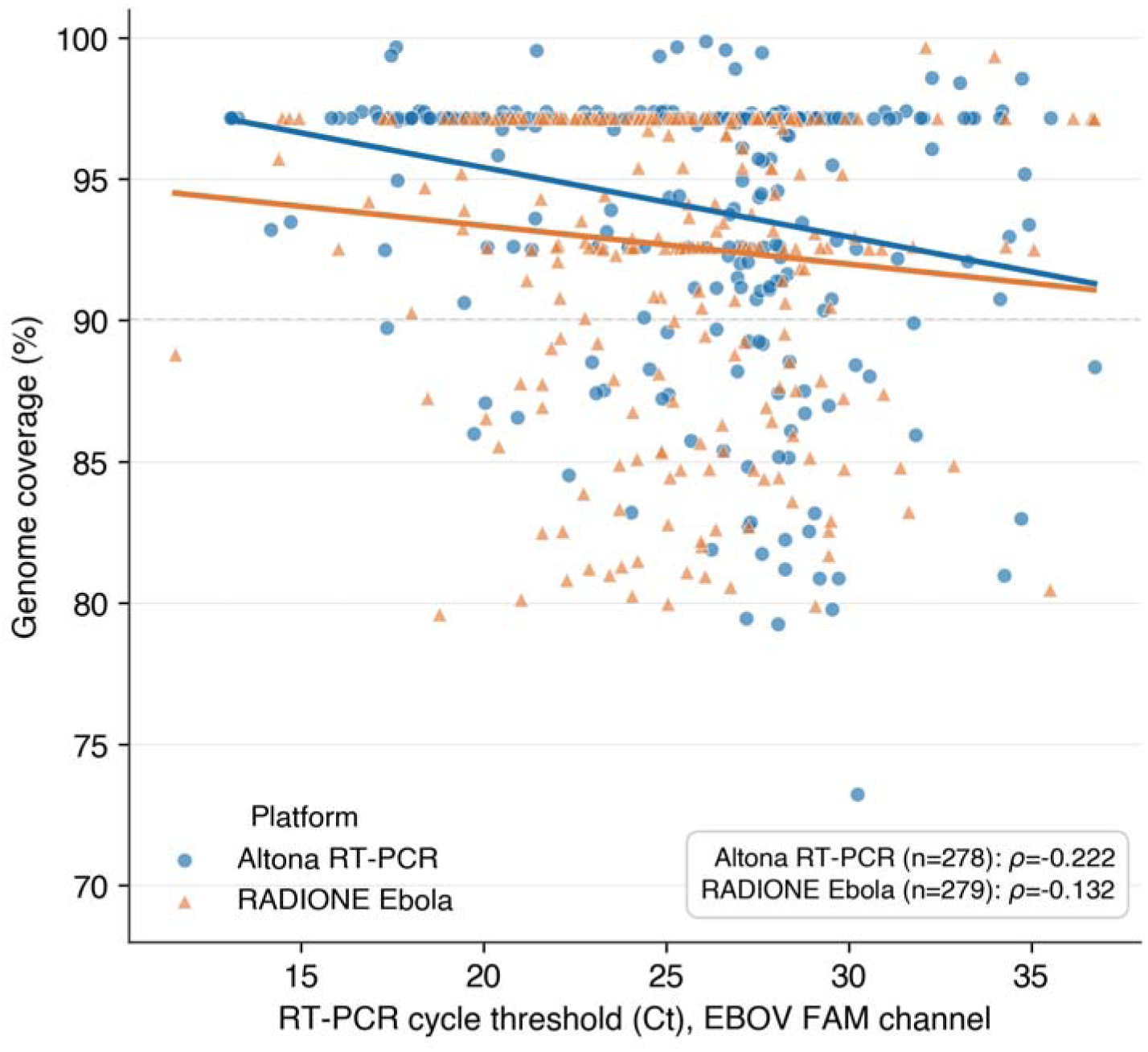
RT-PCR cycle-threshold (Ct) value, by platform, against genome coverage. Each point is one sample; lines are ordinary least-squares fits. Ct is the EBOV FAM-channel value. Higher Ct (lower viral load) was associated with modestly lower coverage, more so for Altona (Spearman ρ=-0.222, n=278) than for RADIONE (ρ=-0.132, n=279); the two coefficients differ (they are not the same value). Dashed line marks 90% coverage.

**Figure S3.**
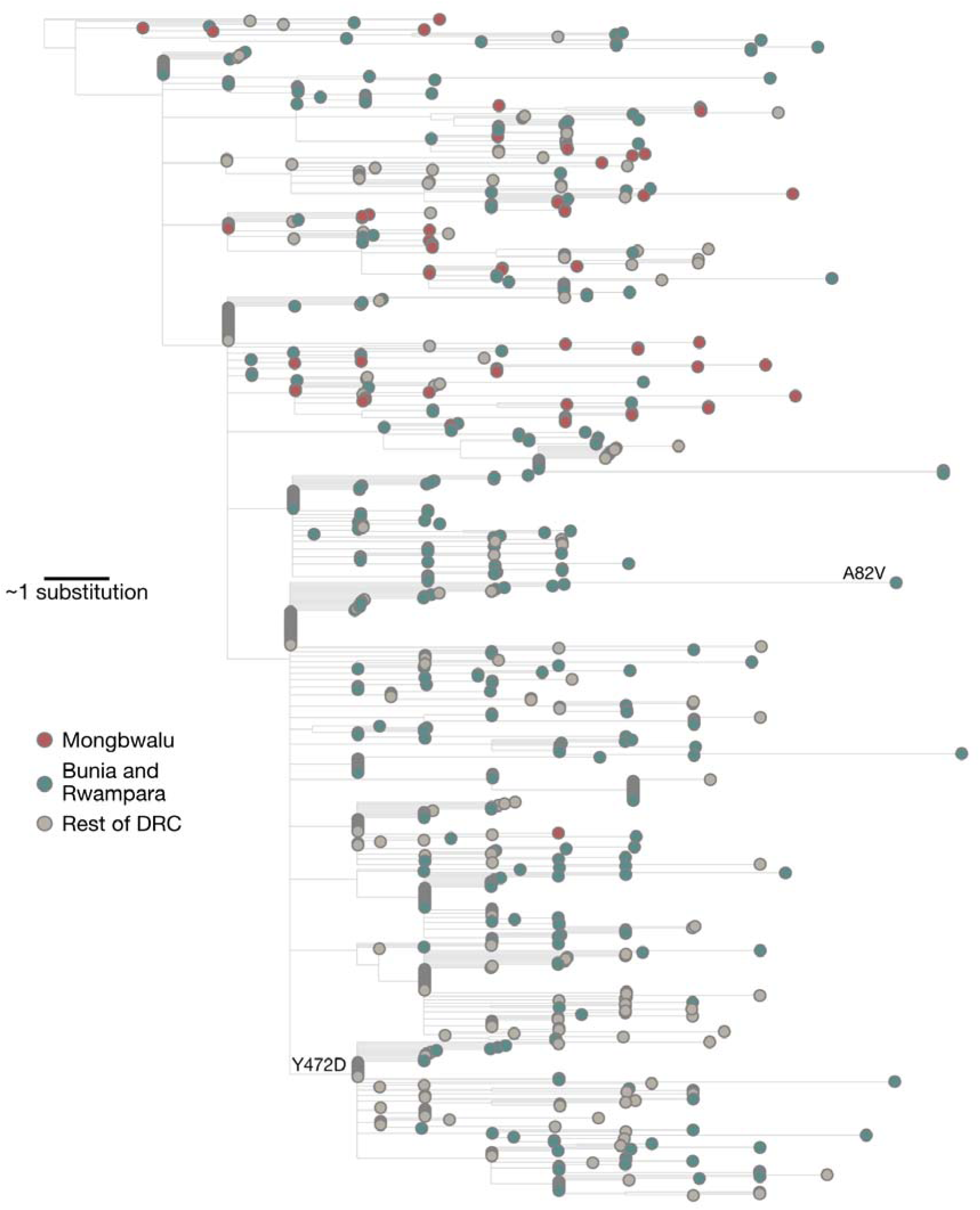
Maximum likelihood phylogeny of the 2026 BVD outbreak in eastern DRC. Substitutions Y472D in NP and A82V in GP are annotated on their respective branches of emergence.

**Figure S4.**
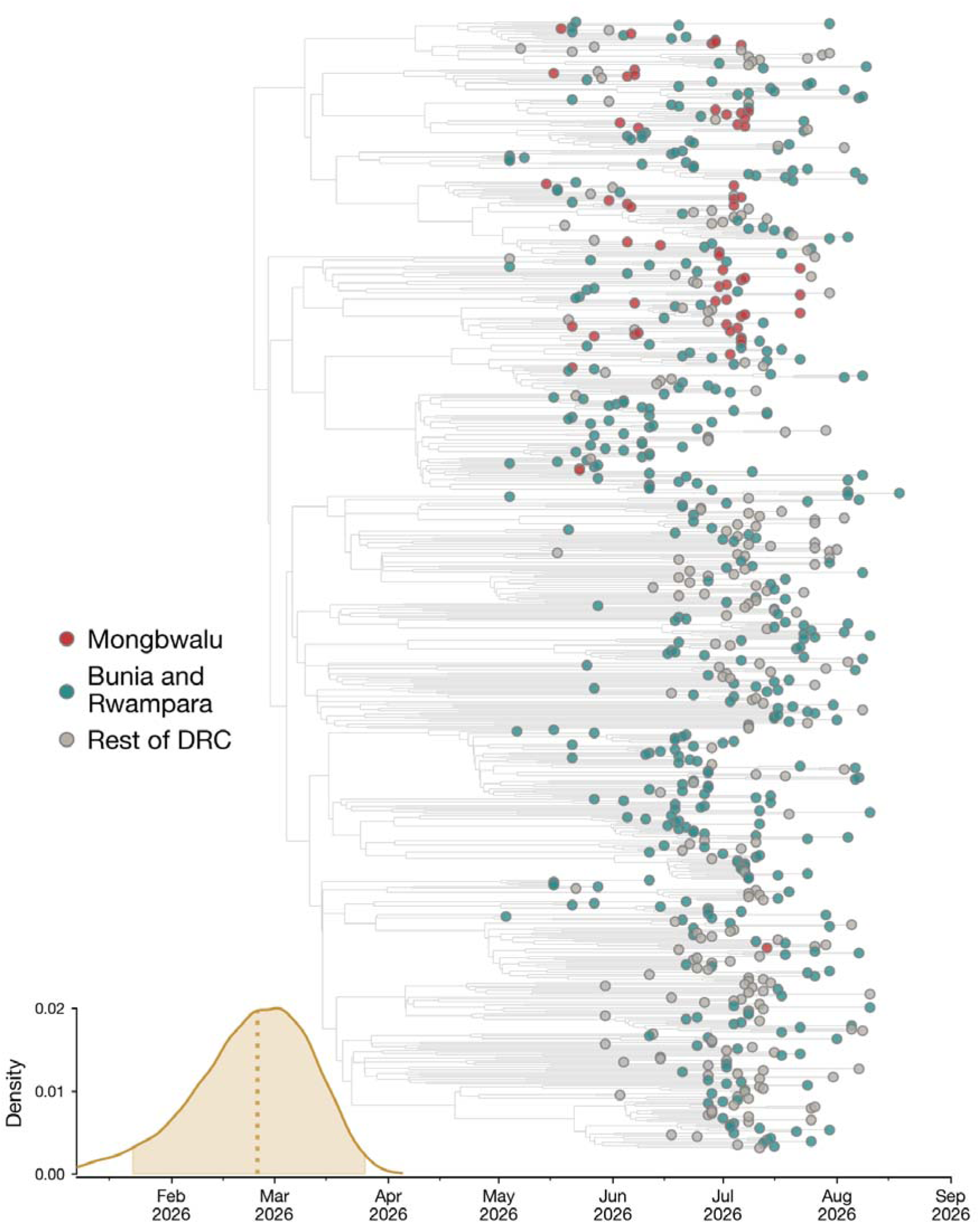
Time-resolved phylogeny of the 2026 BDBV outbreak in the DRC. The density plot shows the time of the most recent common ancestor (tMRCA), with the dotted line indicating the median estimate and the shaded region representing the 95% HPDI.

**Figure S5.**
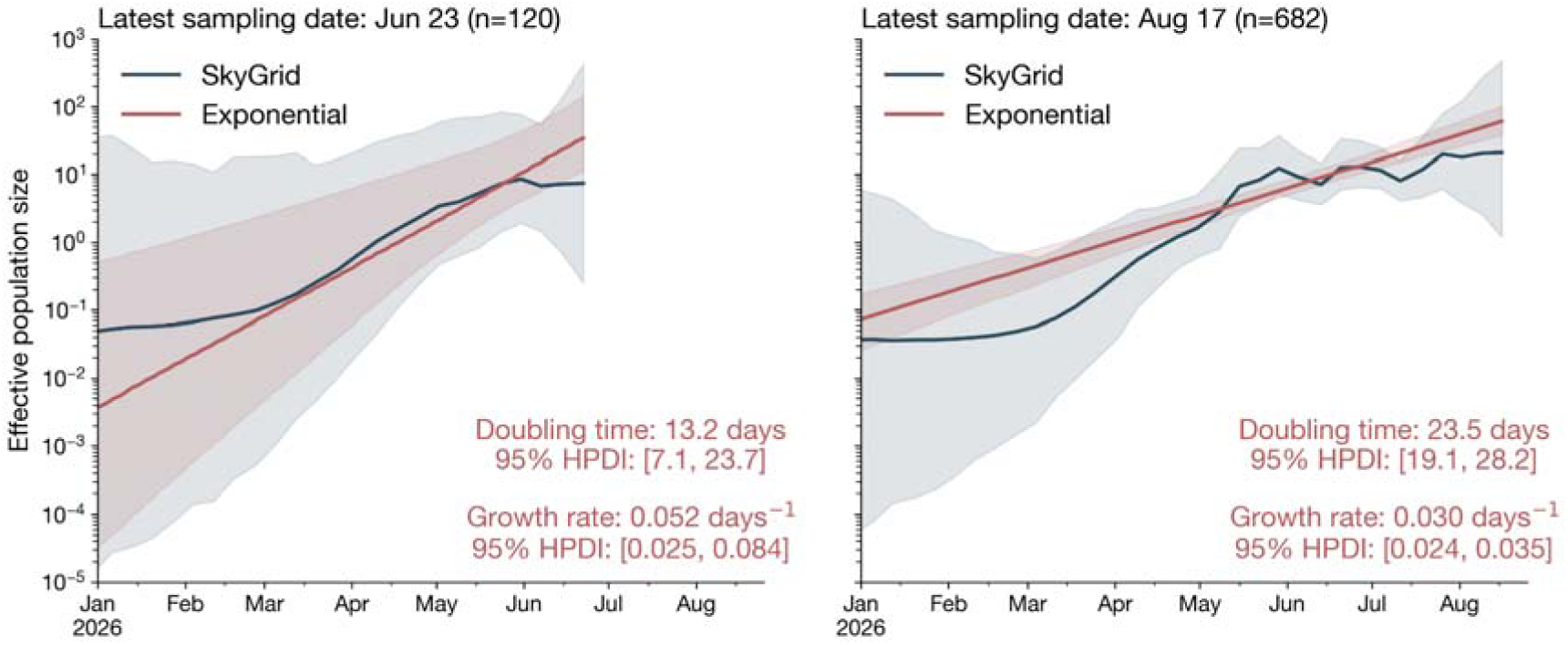
Comparison of exponential growth and SkyGrid coalescent models using the earliest release subsample (left; n=120, sampled through 23 June 2026) and the full dataset (right; n=682, sampled through 17 August 2026). The solid line shows the median estimate and the shaded region represents the 95% HPDI. Colours distinguish the coalescent model. The doubling time and growth rate from the exponential growth model are annotated on each panel.

**Figure S6.**
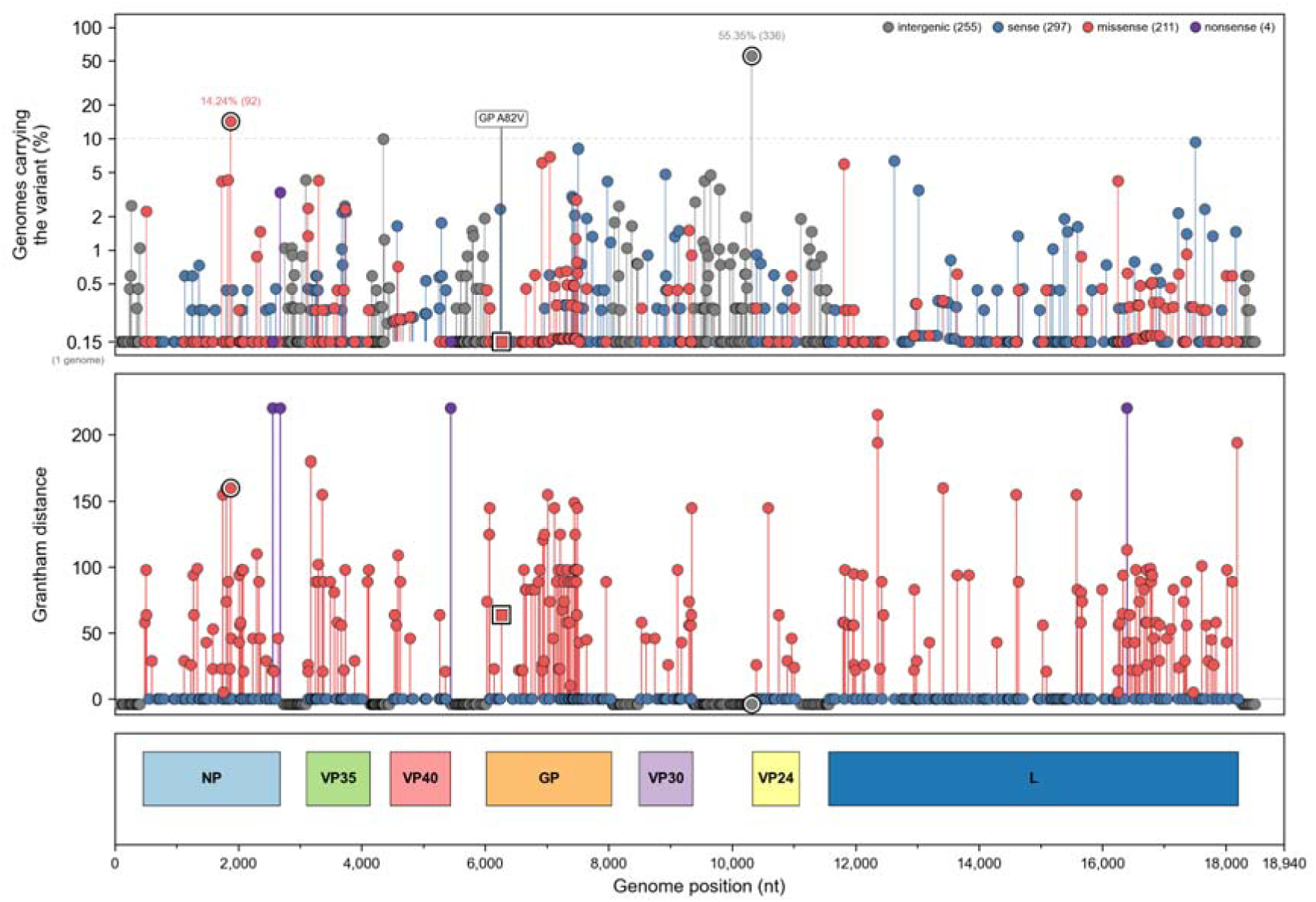
Mutational profile of the 2026 BDBV genomes with nucleotides coordinates relative to BDBV reference genome (GenBank accession: NC_014373.1). The top panel shows the prevalence of each nucleotide substitution across the 682 outbreak genomes (log scale), colored by class (intergenic, sense, missense or nonsense). Sites present in >10% of genomes are marked with a white halo, and GP A82V is highlighted as a square. The middle panel shows Grantham distances for missense and nonsense changes as a measure of their physicochemical impact (higher values indicate more radical amino-acid replacements). The bottom panel shows the BDBDV coding genes along the reference genome axis. All mutations were called relative to the Bayesian-inferred root ancestral sequence.

## Supplementary tables

**Table S1.** Age and sex distribution of sequenced samples.

| Age group — yr | Male<br>no. (%) | Female<br>no. (%) | Total<br>no. (%) |
| --- | --- | --- | --- |
| <5 | 28 (10.1) | 20 (6.2) | 48 (8.0) |
| 5–17 | 45 (16.2) | 45 (14.0) | 90 (15.0) |
| 18–29 | 74 (26.6) | 96 (29.8) | 170 (28.3) |
| 30–44 | 77 (27.7) | 103 (32.0) | 180 (30.0) |
| 45–59 | 32 (11.5) | 38 (11.8) | 70 (11.7) |
| ≥ 60 | 22 (7.9) | 20 (6.2) | 42 (7.0) |
| **Total with age and sex** | **278 (100.0)** | **322 (100.0)** | **600 (100.0)** |
| *Median age (IQR) — yr* | *27 (16–40)* | *30 (20–39)* | *28 (18–39)* |

Percentages are column percentages. Of 686 samples, 600 had both age and sex recorded; 86 were missing age and/or sex (age missing, 84 [12.2%]; sex missing, 32 [4.7%]). Six infants recorded in days or months were included as fractional years.

**Table S2.**
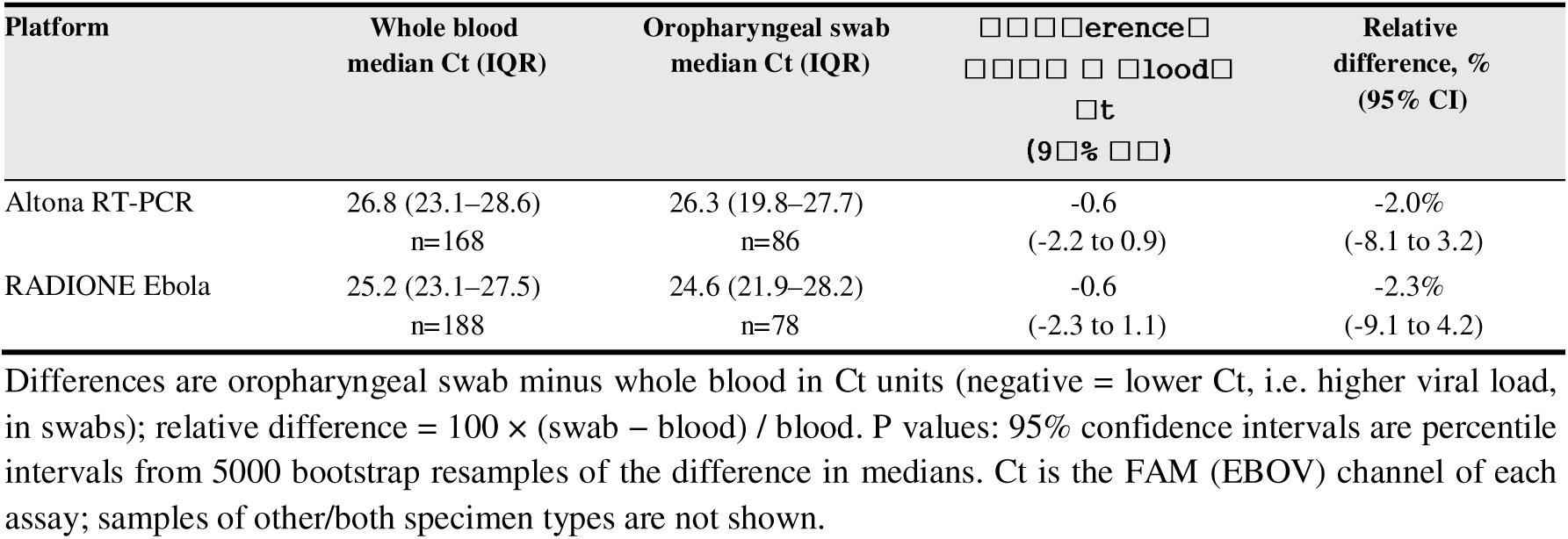
RT-PCR cycle-threshold (Ct) value by specimen type, by platform.

**Table S3.**
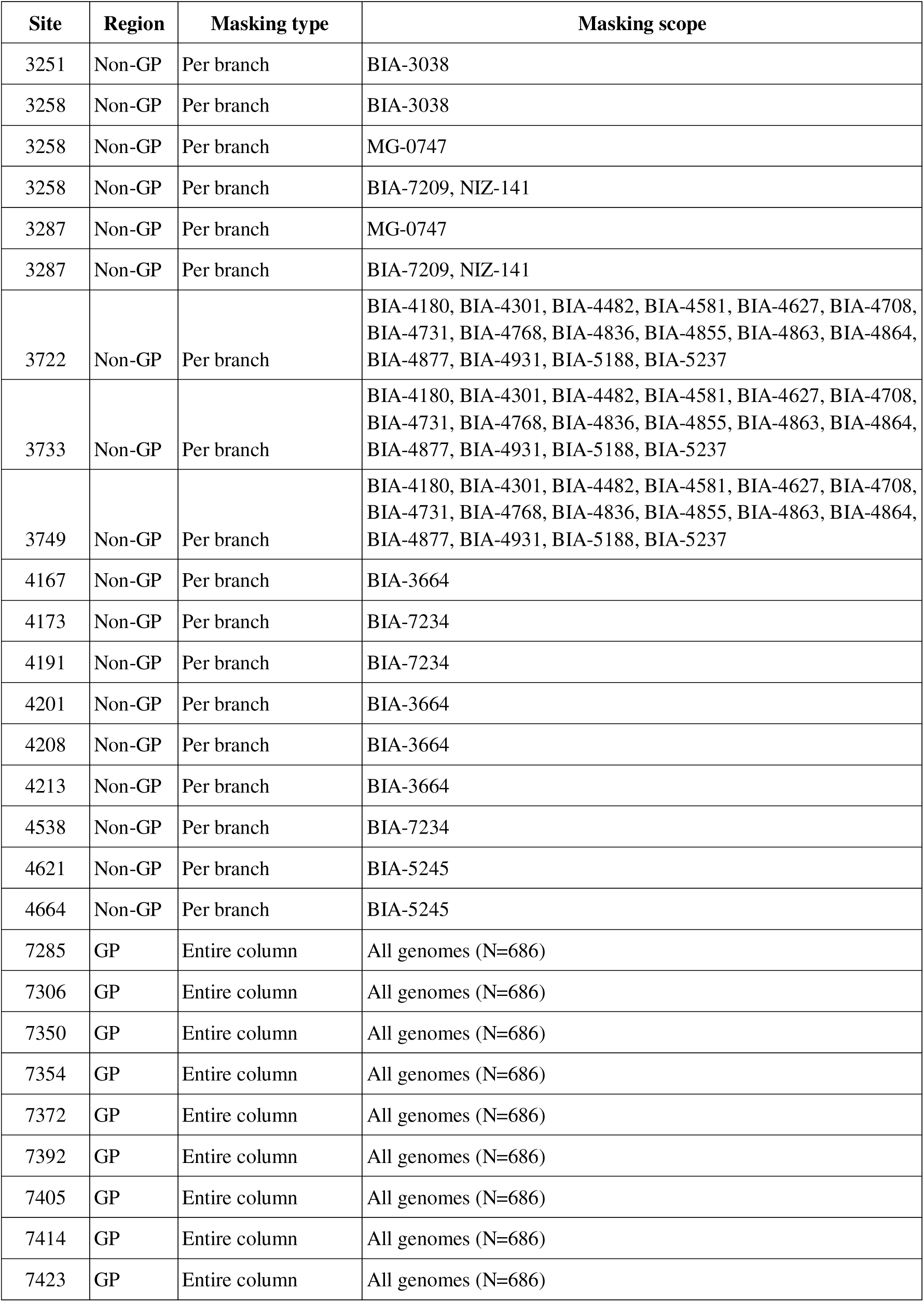
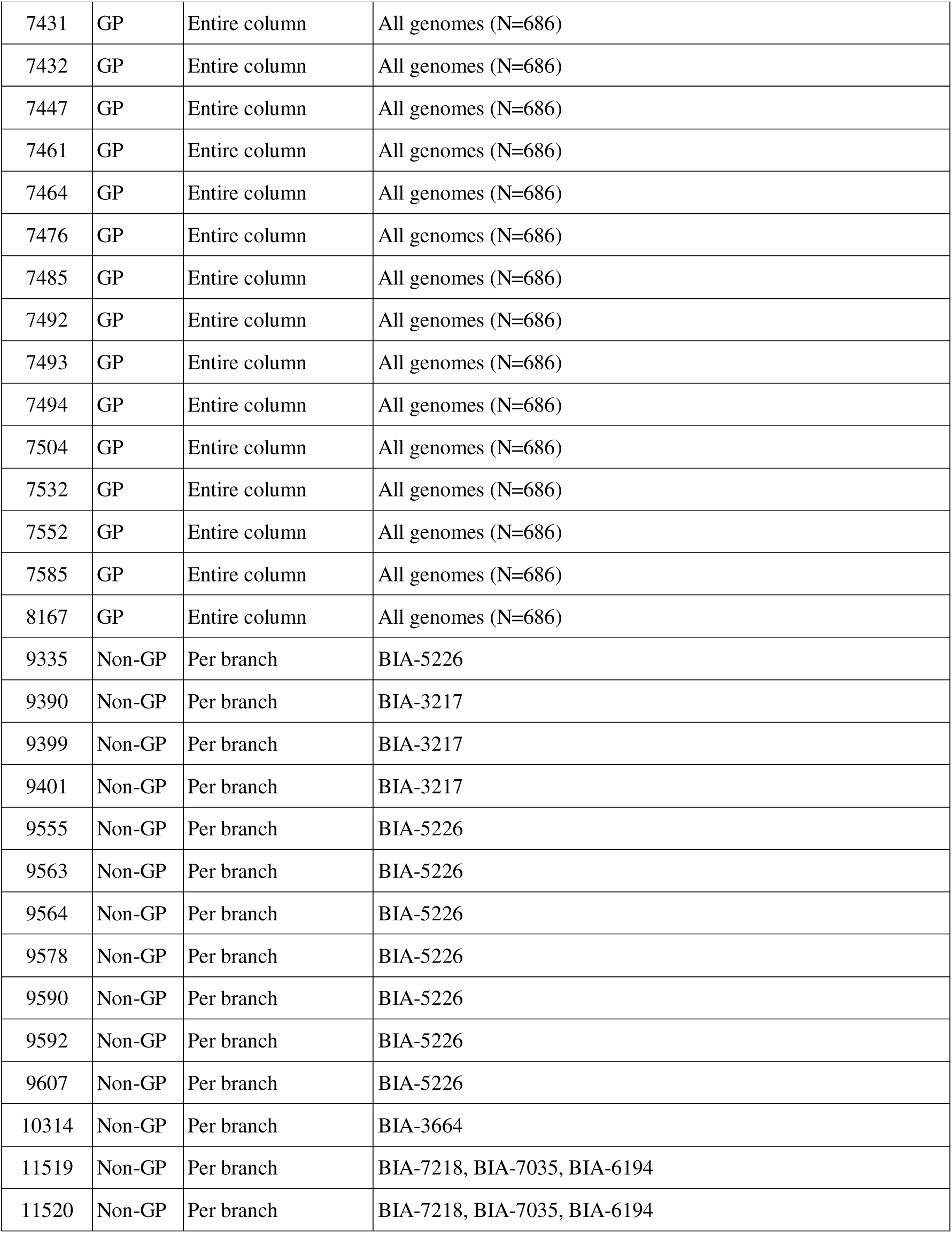
Sites masked due to evidence of ADAR editing. Sites within the GP region were masked across all genomes; sites outside the GP region were masked in affected genomes only. A subset of sites with ADAR editing-type mutations were identified on multiple branches and therefore appear more than once in the table.

