## Supplementary figures and images for "Temporal and Phylodynamic analyses of the 2026 Bundibugyo virus disease outbreak in the Democratic Republic of the Congo"

### Figure S3. Maximum likelihood phylogeny of the 2026 BVD outbreak in eastern DRC.

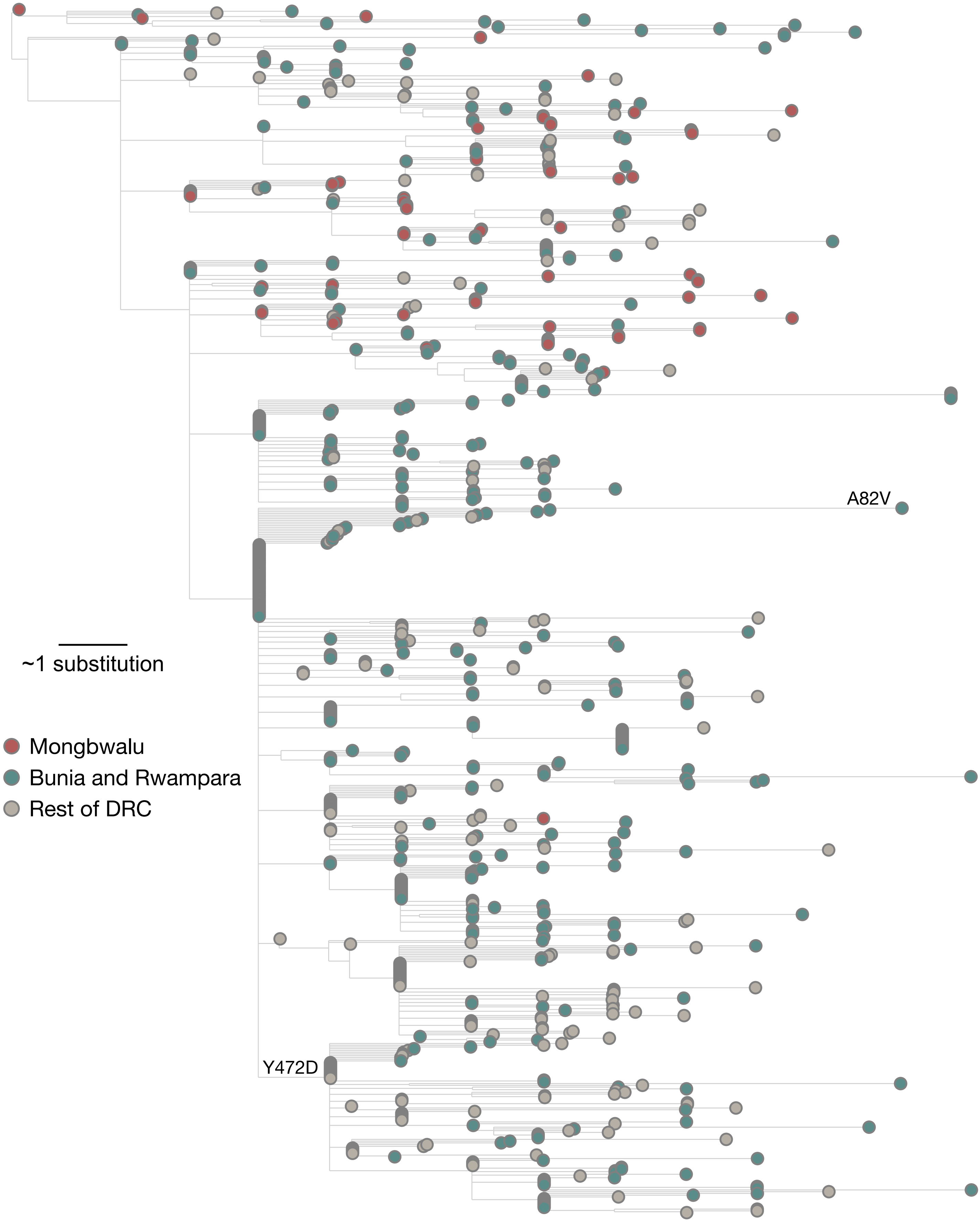

### Figure S4. Time-resolved phylogeny of the 2026 BDBV outbreak in the DRC.

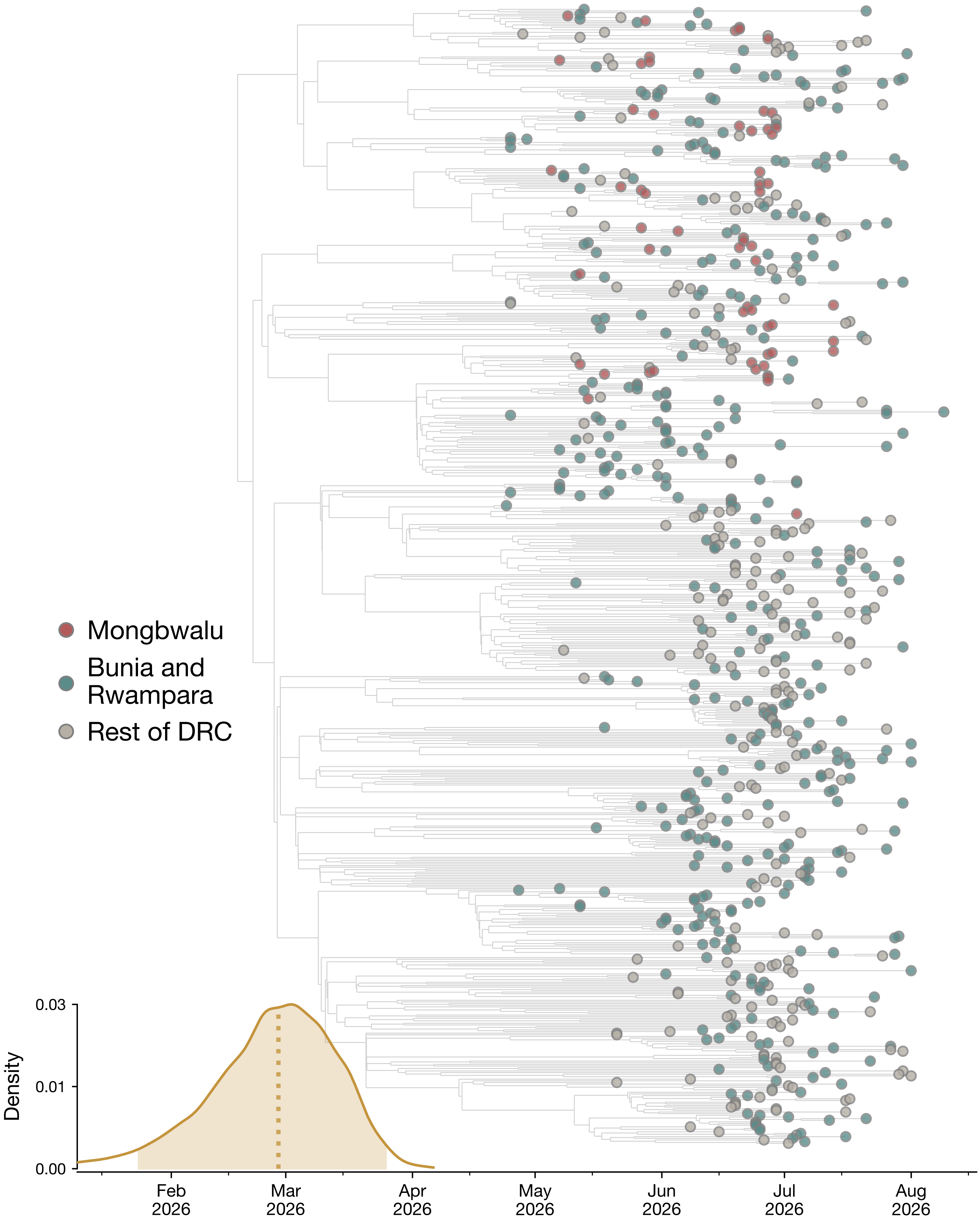

### Figure S5. Comparison of exponential growth and SkyGrid coalescent models using the earliest release subsample (left; n=120, sampled through 23 June 2

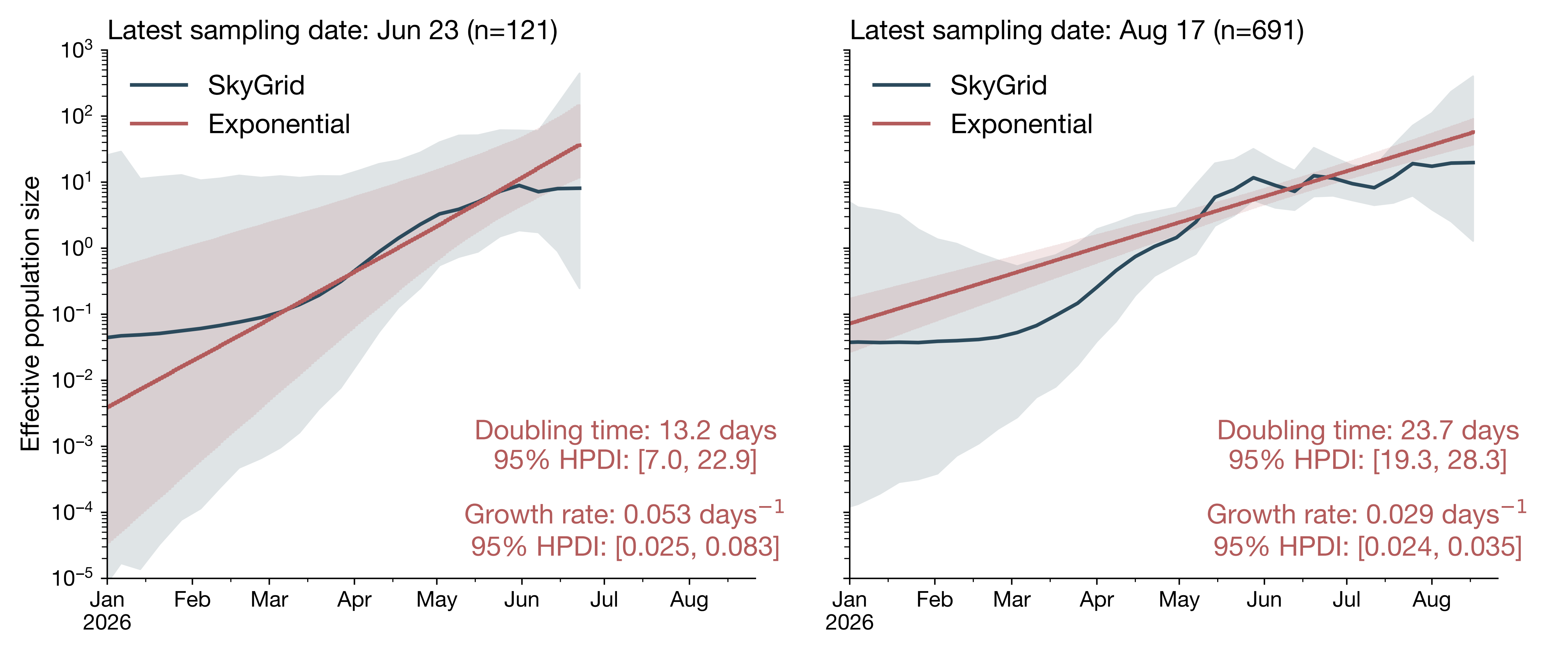
